# Efficacy and safety of therapeutic HPV vaccines to treat CIN 2/CIN 3 lesions: a systematic review and meta-analysis of phase II/III clinical trials

**DOI:** 10.1101/2022.11.11.22282221

**Authors:** Ahmadaye Ibrahim Khalil, Li Zhang, Richard Muwonge, Catherine Sauvaget, Partha Basu

**Affiliations:** Early Detection, Prevention and Infections Branch, International Agency for Research on Cancer (IARC/WHO), Lyon, France

## Abstract

**Objectives:** We aims to assess the efficacy and safety of therapeutic HPV vaccines to treat cervical intraepithelial neoplasia of grade 2 or 3 (CIN2/3).

**Design:** This study is a systematic review and meta-regression that follows the Preferred Reporting Items for Systematic Reviews and Meta-Analyses recommendations.

**Data sources:** PubMed, Embase, Web of Science, Global Index Medicus and CENTRAL Cochrane were searched up January 31, 2022.

**Eligibility criteria:** Phase II/III studies reporting the efficacy of therapeutic vaccines to achieve regression of CIN2/3 lesions were included.

**Data extraction and synthesis:** Two independent reviewers extracted data, evaluated study quality. A random-effect model was used to pool the proportions of regression and/or HPV clearance.

**Results:** 12 trials met the inclusion criteria. Out of the total 734 women receiving therapeutic HPV vaccine for CIN 2/3, 414 regressed to normal/CIN1 with the overall proportion of regression of 0.54 (95%CI: 0.39, 0.69) for vaccinated group. Correspondingly, 166 women receiving placebo only achieving the pooled normal/CIN1 regression of 0.27 (95%CI: 0.20, 0.34). When only including two-arm studies, the regression proportion of the vaccine group was higher than that of control group (relative risk (RR): 1.52, 95%CI: 1.14, 2.04). Six studies reported the efficacy of the therapeutic vaccines to clear high-risk human papillomavirus (hrHPV) with the pooled proportion of hrHPV clearance of 0.42 (95%CI: 0.32, 0.52) for the vaccine group and 0.17 (95%CI: 0.11, 0.26) for the control group and the RR of 2.03 (95%CI: 1.30, 3.16). Similar results were found regarding HPV16/18 clearance. No significant unsolicited adverse events have been consistently reported.

**Conclusions:** The efficacy of the therapeutic vaccines in the treatment of CIN2/3 was modest. Besides, the implementation issues like feasibility, acceptability, adoption, and cost-effectiveness need to be further studied.

**PROSPERO registration number:** CRD42020189617

**Strengths and limitations:**

- This systematic review and meta-analysist on the clinical efficacy and safety of therapeutic human papillomavirus (HPV) vaccines to treat cervical intraepithelial neoplasia of grade 2 or 3 lesions based on phase II/III trials.
- Notre recherche documentaire a impliqué une recherche approfondie d’essais cliniques, en utilisant un large éventail de termes de recherche et sans limitation de langue, de pays ou de date.
- We had to combine studies that were variable in case selection (e.g., some included CIN 2/3 lesions that were positive for HPV 16/18 only while others included lesions associated with any high-risk HPV types).
- Some of the studies did not have a control arm, which made it difficult to ascertain whether the responses observed were due to natural regression alone.

## Introduction

Human papillomavirus (HPV), a non-encapsulated DNA virus with approximately 8000 base pairs belonging to the family *Papillomaviridae* is the most common sexually transmitted infection worldwide.^1^ Persistent infection with high-risk (oncogenic) HPV types may transform the normal cervical epithelium to cervical cancer precursors (cervical intraepithelial neoplasia or CIN) with variable potentials for progressing to cervical cancer, if untreated.^2^ In addition to cervical cancer, HPV is responsible for a significant proportion of cancers of vulva, vagina, penis, anus and oro-pharynx.^3^ HPV types 16 and 18 are the most oncogenic among the high-risk types, and are associated with nearly 75% of cervical cancers worldwide.^4^

The management of CIN lesions depends on the grade of CIN, lesion characteristics, location of the squamo-columnar junction of the cervix and local resources. In higher resourced settings almost all high-grade lesions (CIN 2/3) are treated with large loop excision of transformation zone (LLETZ); a small fraction requiring cold knife conization (CKC).^5^ Women with low-grade lesion (CIN 1) are advised active surveillance only. In limited resourced settings ablative treatment (cryotherapy or thermal ablation) is widely used to treat both low- and high-grade lesions that occupy less than 75% of cervix and are limited to the ectocervix. Rest of the lesions are treated with LLETZ or CKC. While excisional techniques have significantly higher treatment success rates compared to the ablative techniques, the former are difficult to implement in the low- and middle-income countries (LMICs), significantly more expensive and associated with complications like bleeding, infection and adverse impact on subsequent pregnancies.^6^ The World Health Organization (WHO) recommended ablative or excisional treatment of all women (except those living with HIV) positive on HPV test, which will naturally lead to significant number of over-treatments, resultant harms and high burden on the health systems.^7^ Even in the higher resourced settings frequent follow up of the HPV positive women will inconvenience the women and overburden the screening programmes. In summary, the management of women who either have CIN 2/3 lesions or are high-risk HPV positive without any high-grade lesions is not yet optimized.

Therapeutic vaccines designed to eliminate HPV infected cells through stimulation of cell-mediated immune responses have potentials to be the simpler strategies to effectively treat CIN lesions and permanently eliminate the virus.^8^ Various types of therapeutic HPV vaccines have been studied, which include peptide-based vaccines, protein-based vaccines, viral-vectored vaccines, bacterial-vectored vaccines, DNA-based or cell-based vaccines. Though none of the therapeutic vaccines has yet been approved by the regulatory agencies for clinical use, several have undergone evaluation in phase II or III trials.^9^

We performed a systematic review and meta-analysis of the efficacy and safety of therapeutic vaccines in the treatment of patients with histologically confirmed high-grade cervical intraepithelial neoplasia (CIN 2/3) in phase II or phase III clinical trials.

## Methods

### Search strategy

We searched PubMed, Embase, Web of Science, Global Index Medicus and CENTRAL Cochrane to identify phase II/III clinical trials reporting the efficacy of therapeutic HPV vaccines to treat high-grade cervical intraepithelial neoplasia (CIN 2/ 3). If the phase of trial was not specified in the manuscript we used the National Institute of Health (NIH), USA definitions of phase II or III clinical trials available at https://grants.nih.gov/policy/clinical-trials/glossary-ct.htm. All databases were searched from inception till January 31, 2022, without any language restriction. The clinical trials registry platform ClinicalTrials.gov was also searched for any ongoing trial publishing efficacy data. The details of the search strategy for each of the databases are described in the appendix (Table S1). The reference lists of key publications were also scanned. The search strategies combined Medical Subject Headings (MeSH) or equivalent terms in title or abstract regarding HPV and therapeutic vaccine. Only original studies were included. The systematic review was registered with PROSPERO (CRD42022307418).

### Eligibility criteria and Study selection

Two authors (AIK and LZ) independently filtered the title and abstracts and performed the full-text review of the selected articles. Discrepancies were resolved by consensus or in consultation with a senior author (PB). Studies were eligible for inclusion if they reported at least the efficacy outcomes of phase II or phase III trials and participants received a therapeutic HPV vaccine for the treatment of histology confirmed CIN 2/3. Studies designed to evaluate only safety and side effects of the investigational product were not included. The assessment of lesion regression had to be based on cervical histopathology or a combination of cytology, HPV test and colposcopy. Original articles, including randomized control trials, single-arm trials, and case control studies were considered for inclusion.

### Data extraction

Following data were extracted from each study into a Microsoft Excel spread sheet: author and year of publication, study period, study phase, study design, country where conducted, study population and their age, interventions for vaccinated (vaccination protocol) and control (if applicable) groups, follow-up interval and number of women followed up and their outcomes. The primary outcome for our systematic review was the regression of histology-confirmed CIN 2/3, defined as documentation of a normal or a CIN 1 histology result or a combination of negative high-risk HPV test and absence of high-grade cervical lesion on cytology or colposcopy (when cytology was abnormal but the abnormality was lower than high-grade) at follow up. The secondary outcomes were high-risk HPV clearance and safety.

### Quality assessment of studies

The methodological quality of each study included in the review was independently assessed by two authors (AIK, LZ). We used the study quality assessment tool of the National Heart, Lung, and Blood Institute (NHLBI) to categorize quality of the studies as good, fair, or poor using the standardised checklist of the panel for systematic reviews.^6,10,11^ Discrepancies were discussed until consensus was reached, with assistance from the senior author (PB), if necessary.

### Data analysis

The primary endpoint was regression of CIN 2/3 at follow-up based on the criteria described earlier. The proportion of cases undergoing regression in the vaccine group was compared to that in the control group using placebo in a meta-analysis. Analyses were stratified by median follow-up intervals (≤6 months vs. >6 months) and mode of delivery of the vaccine (oral vs. injectable). All meta-analyses were conducted using the “meta” and “metafor” R package. The I^2^ statistic was used to assess rather the proportion of total variability due to between-study variability. A random effects model was adopted regardless of the extent of heterogeneity as evidence has shown that it is more robust compared to fixed effect models.^12,13^ The combined efficacy estimated as proportion of patients having regression together with their 95% confidence intervals (CI) were presented in forest plots. Besides, the relative risk (RR) with corresponding 95% CI was evaluated based on two-arm studies with the control arm of placebo. The R software (version of 4.1.0) was used for all the statistical analysis.

## Results

### Study selection

We identified 202 articles, among which, 12 studies published between 2004 and 2021 met the selection criteria (Figure 1). The included studies represented six randomized controlled trials (RCTs) (Ikeda et al 2021; Harper et al 2019; Trimble et al 2015, Kaufman et al 2007 and Garcia et al 2004 ; Choi et al 2020);^14-19^ three case-control studies (Rosales et al 2014 and Garcia-Hernandez et al 2006, Corona Gutierrez et al 2004)^20-22^ and three single-arm trials (Park et al 2019; Brun et al 2011 and Einstein et al 2007).^23-25^ All the included studies recruited women with histology confirmed diagnosis of CIN 2/3. Characteristics of the included studies are listed in Table 1. Five studies administered placebo to the participants belonging to the control group; three studies treated the control participants with excision or ablation (the standard-of-care treatment for CIN 2/3) that are not included in our analysis. In the RCT by Choi et al the vaccine was administered to all participants, but at different doses (4 mg or 1 mg) in two randomized arms. All the subjects in the study were analysed together as recipients of therapeutic vaccine. Rosales et al recruited a total of 300 subjects with high grade lesions to be treated with therapeutic vaccine; 112 of them were recurrent cases after ablative or excisional treatment. The recurrent cases were excluded from the efficacy analysis. Among the 12 studies included, 4 studies were categorized as good (33%) 8 studies were categorized as fair (67%).

**Figure 1.**
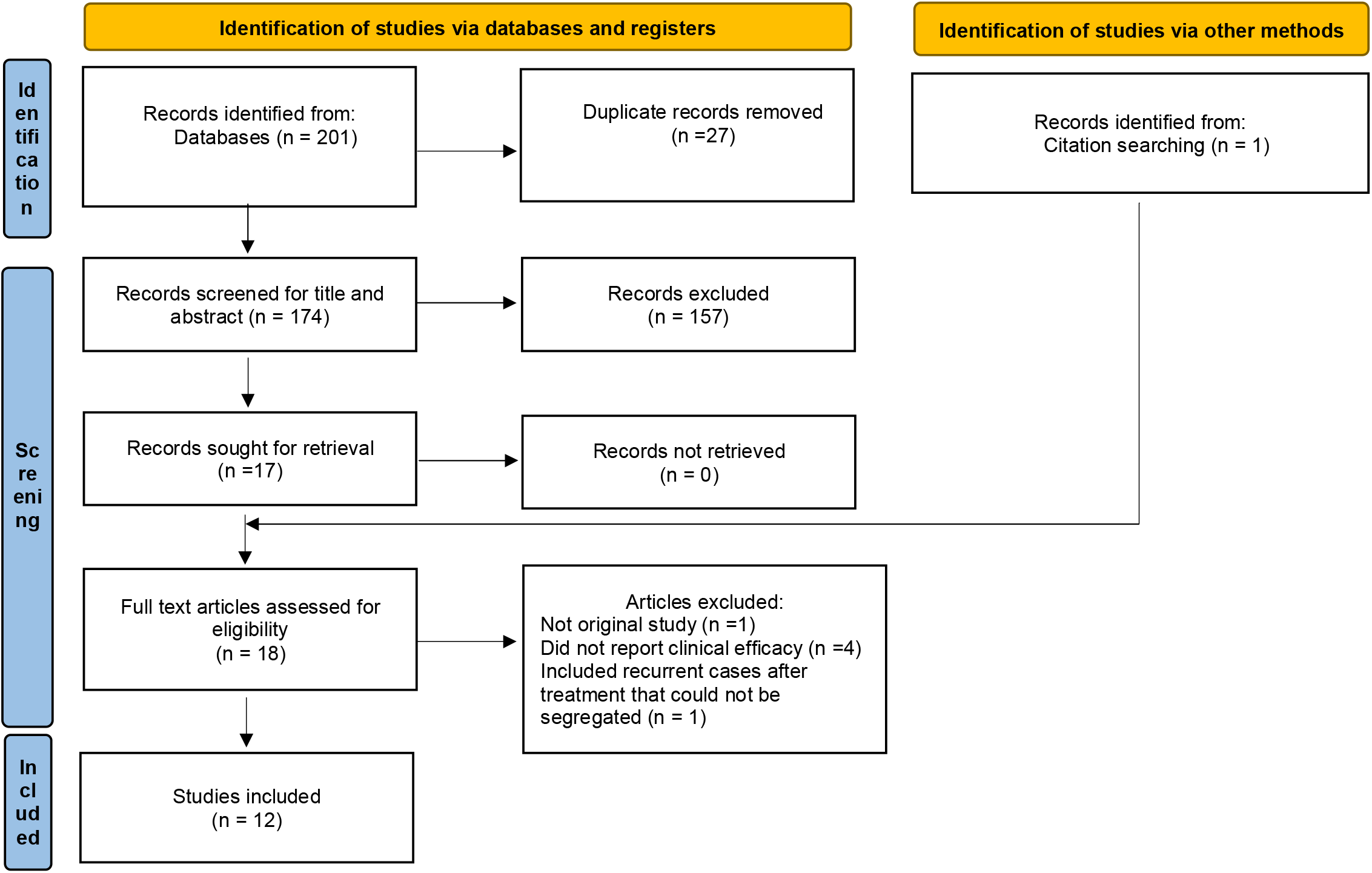
PRISMA flow diagram of study selection process

**Table 1.**
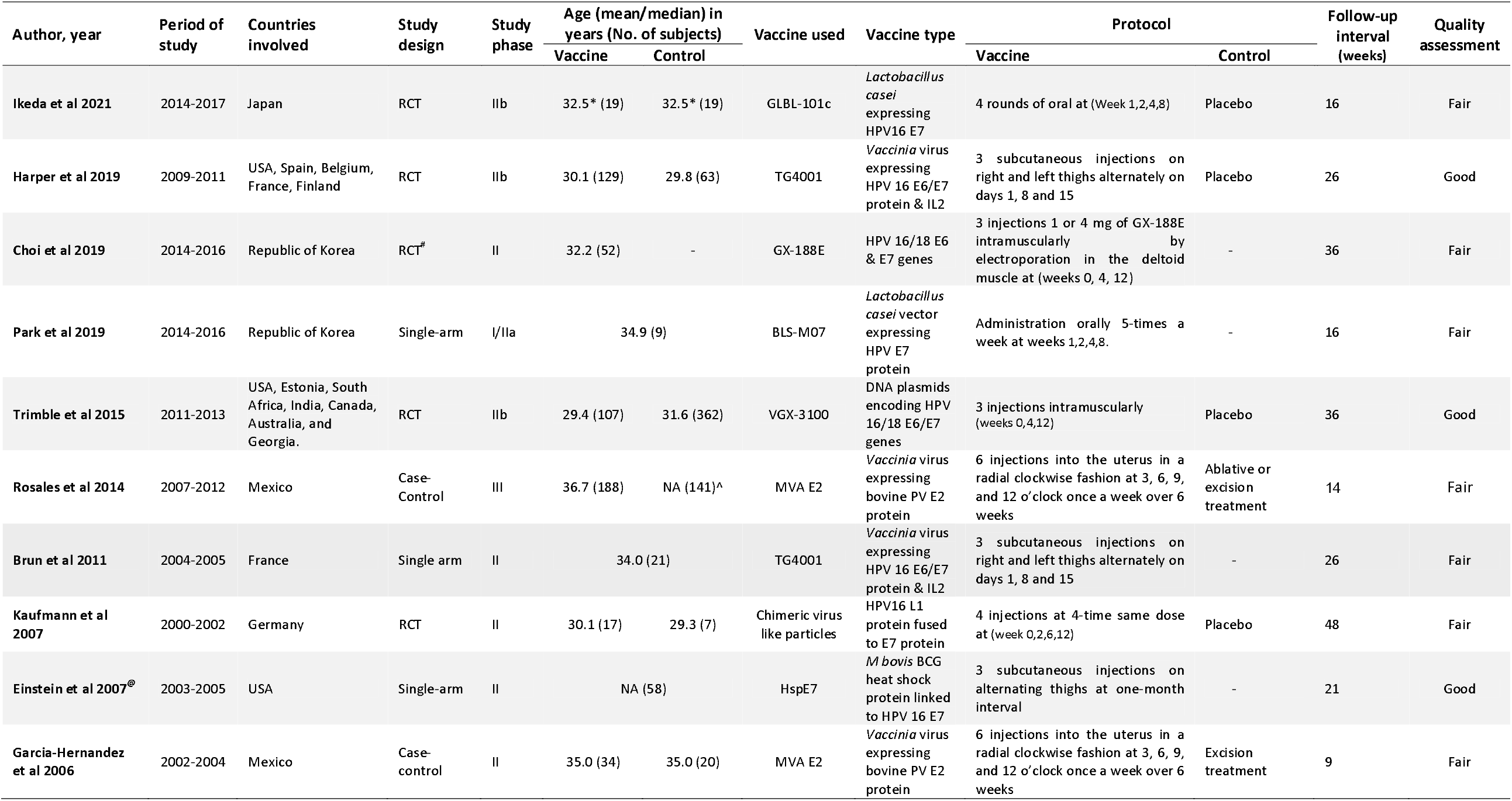
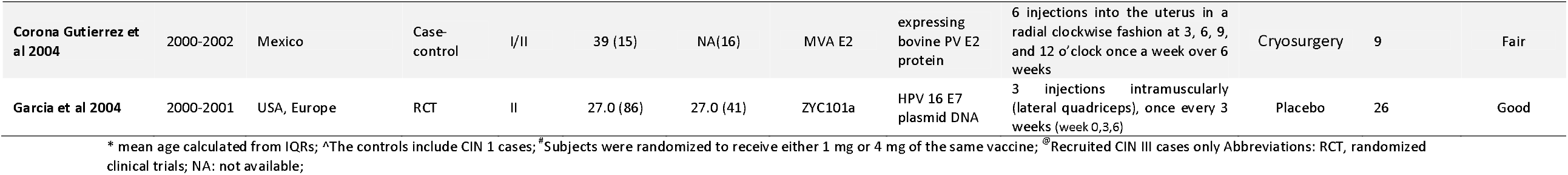
Characteristics of the included studies

### Primary outcomes

The median follow-up interval was 26 weeks (ranging from 9 to 48 weeks). All studies reported regression of CIN 2/3 among the vaccine recipients. In the single-arm study by Brun et al the participants were followed with cervical cytology and high-risk HPV test. Women positive on either test had colposcopy and biopsy. Any woman having histopathology proved CIN 2/3 or a positive HPV test result or a high-grade abnormality on cytology or colposcopy at follow up was considered as treatment failure. Rest of the studies performed cervical biopsies for all the participants and the outcome was based on histopathology report. Out of the total 734 women receiving therapeutic HPV vaccine for CIN 2/3, 414 regressed to normal/CIN 1 (normal: 253; CIN 1: 52; normal/CIN 1 unspecified: 109) at follow-up. The pooled proportion of histopathologic regression to normal/CIN 1 was 0.54 (95%CI: 0.39, 0.69) for vaccinated group at variable follow-up intervals. Among the total 166 women receiving placebo only the proportion achieving regression was 0.27 (95%CI: 0.20, 0.34) (Figure 2A). The proportion of regression to normal/CIN 1 in the vaccine group was significantly higher than that of control group (RR: 1.52, 95%CI: 1.14, 2.04) (Figure 2B).

**Figure 2:**
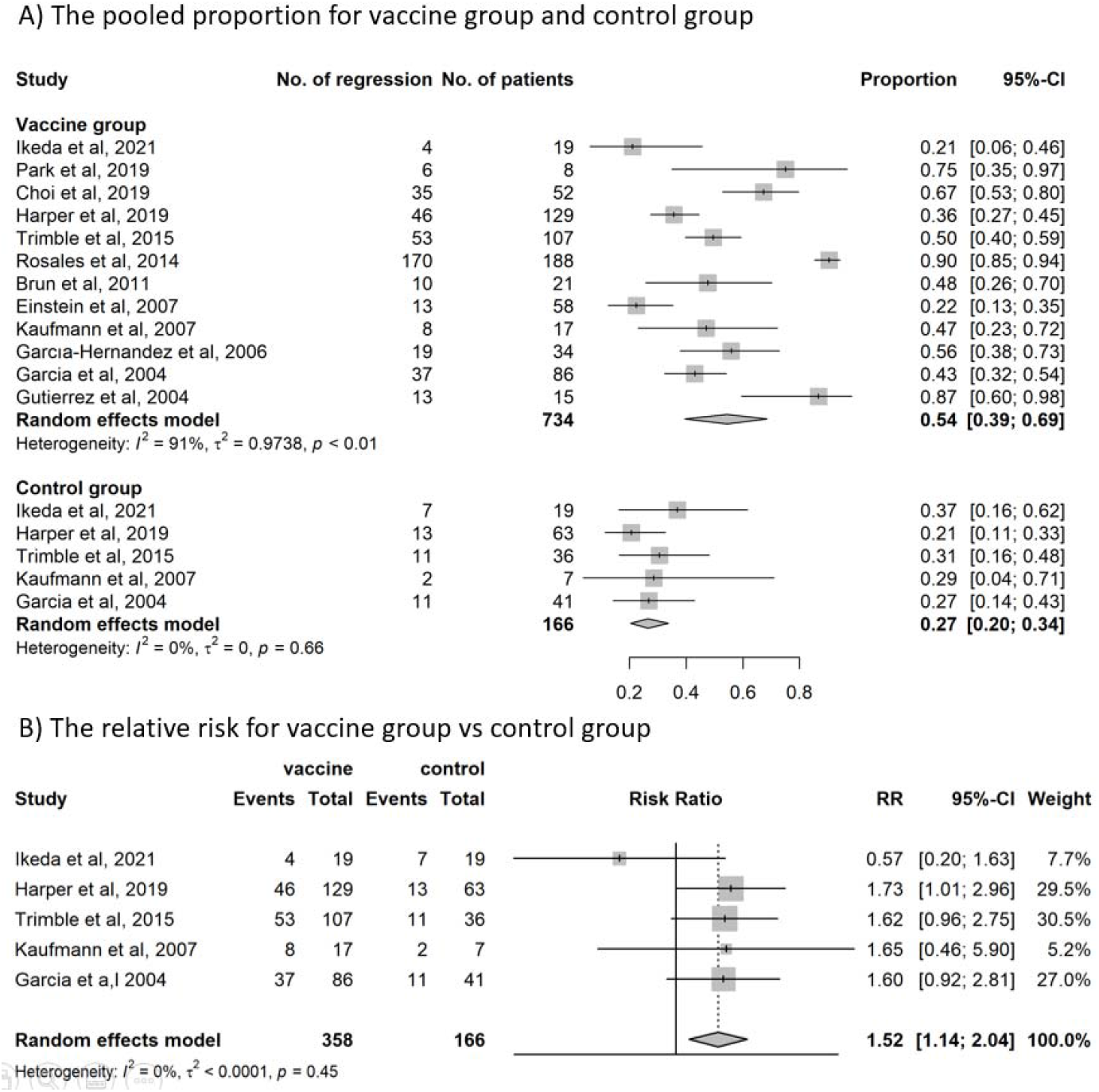
CIN 2/3 lesions regressing to normal or CIN1; 2A) The pooled proportion or lesions regressing in vaccinated and control groups; 2B) The relative risk for vaccinated group vs control group

The heterogeneity between the studies was high for the vaccine group (I^2^ = 92%). The proportion of vaccinated subjects achieving regression was lower compare to those assessed after follow-up exceeding 6 months [0.48 (95%CI: 0.39, 0.56)] compared to those assessed within 6 months [0.62 (95%CI: 0.33, 0.84)] (Figure S1). No difference in efficacy was observed between oral administration [proportion regressing 0.44 (95%CI: 0.12, 0.82)] and intramuscular or intradermal administration [proportion regressing 0.54 (95%CI: 0.38, 0.70)] (Figure S2). The proportion of regression to normal/CIN 1 in the vaccine group was significantly higher than that of control group (RR: 1.52, 95%CI: 1.14, 2.04) (Figure 2B).

### Secondary outcomes

Efficacy of the therapeutic vaccines to clear high-risk HPV was reported by six studies that included a total of 357 women; 145 of them cleared infection at post-treatment follow up. Some of these studies evaluated the clearance of HPV 16 and 18 only.^16,19,24^ The pooled proportion of women having high-risk HPV clearance was 0.42 (95%CI: 0.32, 0.52) in the vaccine group and 0.17 (95%CI: 0.11, 0.26) in the control group receiving placebo only (Figure 3A). Only three studies evaluating high-risk HPV clearance included a control arm (Harper et al 2019, Trimble et al 2015, Kaufmann et al 2007) Proportion of HPV clearance was significantly higher in the vaccinated group compared to that in the control group (RR: 2.03 (95%CI: 1.30, 3.16) (Figure 3B).

**Figure 3:**
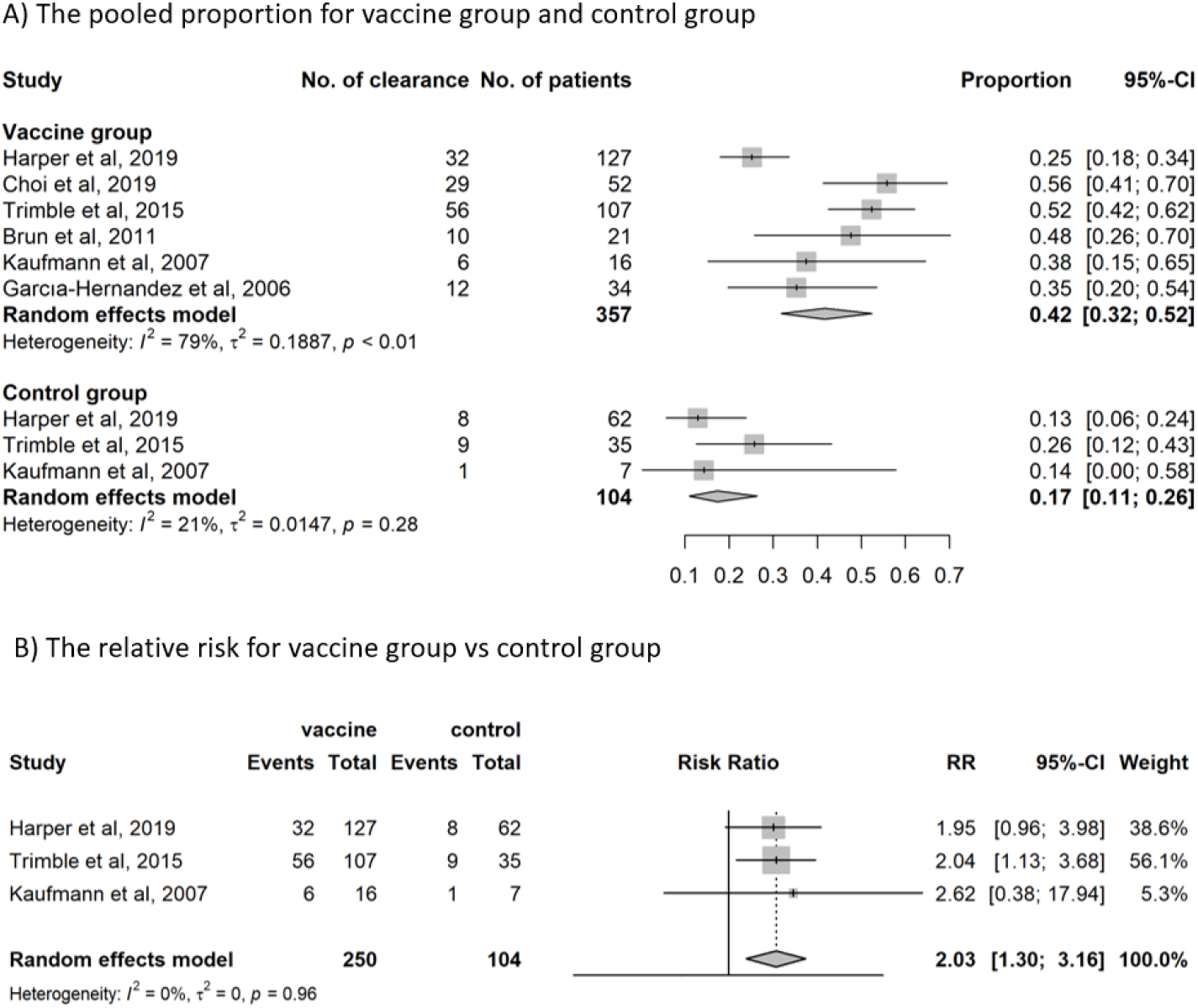
High risk HPV clearance; 3A) The pooled proportion of cases having HPV clearance in vaccinated and control groups; 3B) The relative risk of HPV clearance for vaccinated group vs control group

HPV16/18 clearance was observed in 87 of the 199 women with CIN 2/3 receiving the therapeutic vaccine. The pooled proportion was 0.41 (95%CI: 0.30, 0.54) in the vaccinated group, and 0.21 (95%CI: 0.13, 0.32) in the control group (Figure 4A). The proportion of HPV16/18 clearance was significantly higher following vaccination compared to those receiving placebo only (RR: 2.00 (95%CI: 1.22, 3.27)) (Figure 4B).

**Figure 4:**
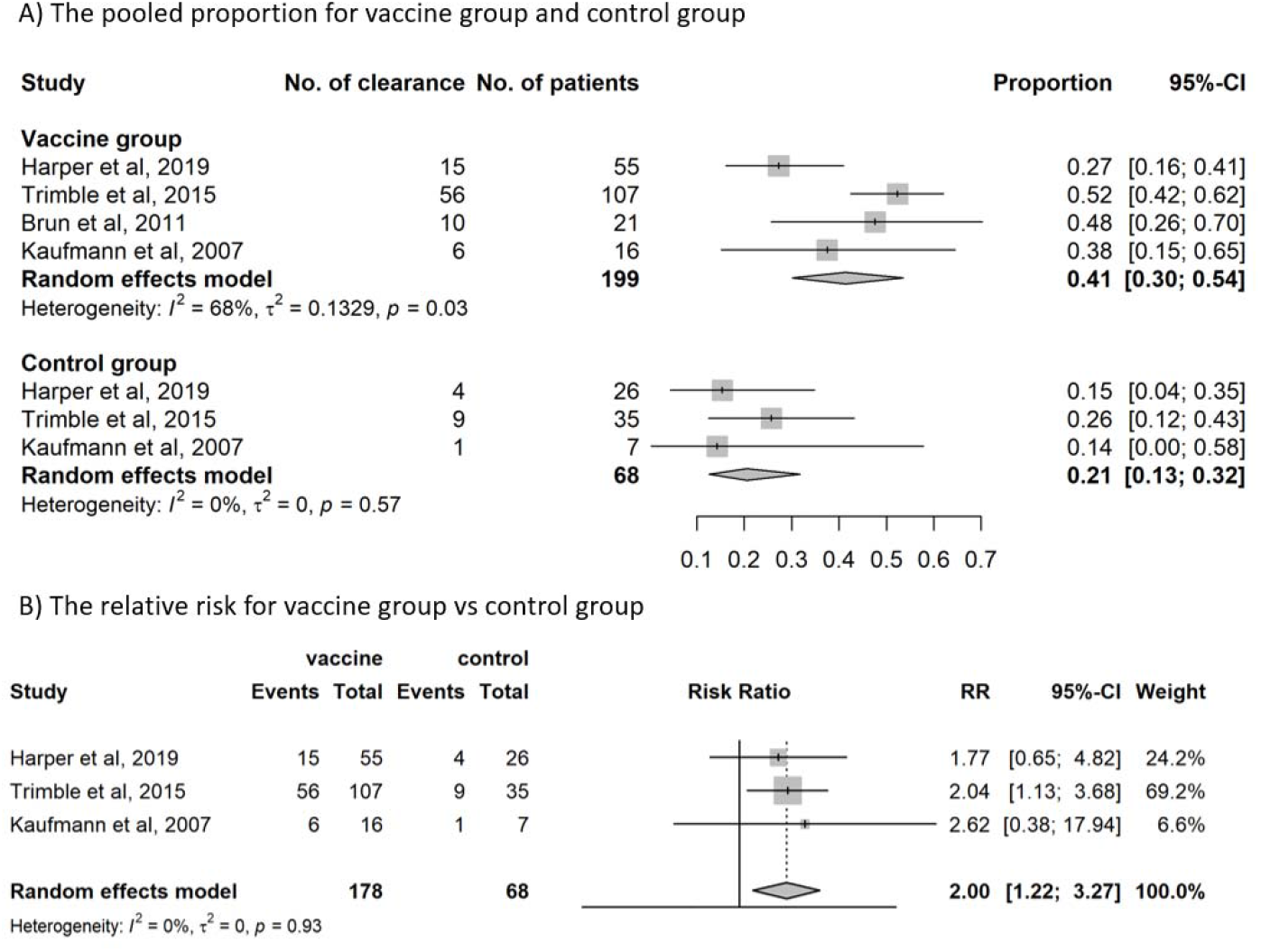
HPV 16/18 clearance; 3A) The pooled proportion of cases having HPV 16/18 clearance in vaccinated and control groups; 3B) The relative risk of HPV 16/18 clearance for vaccinated group vs control group

### Adverse events

We assessed adverse events of therapeutic vaccines from the five studies reporting such events for vaccinated and placebo groups. Only two serious adverse events related to the vaccines have been reported - one case of severe lymphadenopathy reported by Harper et all 2019 et al and one case of severe injection site reaction requiring surgical intervention reported by Trimble et al (Table 2). The solicited adverse events, especially injection site reactions, were higher in the vaccinated group as expected (Table 2). No other significant unexpected adverse events have been consistently reported following therapeutic vaccination by any of the publications.

**Table 2.** Comparison of adverse events reported following administration of therapeutic vaccine and placebo

| Adverse Events |  | Vaccine group<br>(n/N) % | Control group<br>(n/N) % |
| --- | --- | --- | --- |
| **Serious Adverse Events (SAE)** |  |  |  |
| Lymphadenopathy |  | 1/136 (1) | 0/70 (0) |
| Injection-site pain discontinued dosing, to seek surgical resection |  | 1/107 (1) | 0/36 (0) |
| **Adverse Events (AE)** |  |  |  |
| Injection Site |  |  |  |
|  | Hematoma | 5/136 (4) | 6/70 (9) |
|  | pain | 75/111 (68) | 16/50 (32) |
|  | Erythema | 120/236 (50) | 27/92(29) |
|  | Induration | 18/111 (16) | 0/50 (0) |
|  | Tenderness | 104/125 (83) | 32/42 (76) |
|  | Swelling or oedema | 62/125 (49) | 14/42 (33) |
| Headache |  | 94/414 (23) | 37/193 (19) |
| Pain |  |  |  |
|  | General | 114/125 (91) | 38/42 (90) |
|  | Abdominal lower | 7/130 (5) | 6/69 (9) |
|  | Limb | 9/111 (8) | 2/50 (4) |
| Nausea |  | 44/391 (11) | 21/162(13) |
| Vomiting |  | 7/111 (6) | 2/50 (4) |
| Lymphadenopathy |  | 9/136 (7) | 0/70 (0) |
| Fatigue |  | 77/236 (33) | 22/92 (24) |
| Pyrexia >38°C |  | 7/136 (5) | 0/70 (0) |
| Influenza like illness (flu syndrome) |  | 8/159 (5) | 2/82 (2) |
| Diarrhoea |  | 10/111 (9) | 3/50 (6) |
| Myalgia |  | 48/125 (38) | 14/42 (33) |
| Malaise |  | 39/125 (31) | 11/42 (26) |
| Peripheral neuropathy |  | 0/19 (0) | 1/19 (5) |
| Itch |  | 1/19 (5) | 0/19 (0) |
| Skin rash |  | 2/19 (10) | 0/19 (0) |

## Discussion

The present systematic review demonstrates that a wide variety of therapeutic vaccines have been evaluated in phase II/III trials. HPV mediated tumorigenesis necessitates overexpression of the viral proteins E6 and E7 in the cells undergoing malignant transformation.^26^ Since these proteins are not expressed elsewhere in the body, the HPV infected transforming cervical cells expressing the proteins are ideal targets for therapeutic vaccines. Most therapeutic vaccines have E6 and E7 proteins as the target antigens. E2 viral protein is also an attractive candidate for therapeutic vaccines, since the protein is highly expressed before viral genome integration. Other agents are added to the vaccine to potentiate the immune response by decreasing the immune-suppressive elements in the tumour microenvironment (e.g., Interleukin-2). Orally administered therapeutic vaccines using HPV 16 E7 expressing Lactobacillus casei induce gut-associated lymphoid tissue immune response by entering through Peyer’s patches in the small intestine. The oral vaccines are likely to have higher acceptability and are shown to be as effective as injectable vaccines to treat CIN 2/3 lesion in our meta-analysis.

Our systematic review based on 12 fairs to good quality studies demonstrates that the therapeutic vaccines currently available have a modest efficacy in achieving regression of high-grade cervical cancer precursor lesions. While the vaccines achieved an overall regression of only 54% of CIN 2/3 lesions, treatment with standard of care excisional techniques can successfully treat more than 90% of such lesions.^27^ Ablative treatment with either cryotherapy or thermal ablation has lower success rate compared to excision, yet the cure rate exceeds 80%.^6^ The proportion of regression achieved after vaccination was about 50% higher than that observed without any treatment (due to natural clearance), which may not be considered enough to change the current practices of treating the high-grade precursor lesions with ablation or excision. The MVA E2 vaccine containing the bovine papillomavirus E2 protein expressed by modified vaccinia virus Ankara (MVA) demonstrated a 90% regression of CIN 2/3 lesions, the highest reported among all the vaccines.^20^ Since the study did not have any ‘no-treatment’ control arm, the proportion of cases that would have regressed naturally could not be ascertained. Only the vaccine TG4001 (Transgene, Illkirch-Graffenstaden, France) based on MVA expressing HPV16 E6 and E7 proteins along with interleukine-2 had a statistically significant higher proportion compared to ‘no treatment’ (RR 1.73; 95%CI 1.01, 2.96).^15^ Vaccines containing HPV 16/18 E6 or E7 proteins as target antigens could successfully treat the CIN 2/3 lesions associated with any high-risk HPV types, substantiating the cross-protective efficacy of at least some of the therapeutic vaccines.^28^

The modest efficacy of the vaccines observed in achieving clearance of any high-risk HPV or HPV 16/18 alone may have greater public health significance as there is no standard-of-care treatment for high-risk HPV infection. Overall, the therapeutic vaccines cleared 42% of any high-risk HPV infections and 41% of HPV 16/18 infections within the short follow up interval. The clearance proportion in the vaccinated women was double compared to that observed with placebo. The most effective of all the vaccines in achieving viral clearance was VGX 3100 (Inovio Pharmaceuticals, Plymouth Meeting, PA, USA) that comprised of DNA plasmids encoding E6 and E7 genes of HPV 16 and HPV 18.

We need to identify an appropriate niche for the therapeutic vaccines within the cervical cancer prevention continuum. The WHO call to eliminate cervical cancer as a public health problem globally is supported by three strategic pillars – 90% of girls being vaccinated before 15 years of age, 70% of women being screened at least twice by 45 years of age with a high-performance test, and treatment of 90% women with screen detected precancers and cancers.^29^ These targets need to be achieved by the year 2030 to successfully eliminate the cancer by the millennium end. Global access to prophylactic HPV vaccine has been highly inequitable with only 12% of adolescent girls being vaccinated in the LMICs till 2019.^30^ The recent endorsement of a single dose of the vaccine by the WHO is expected to improve the situation in the coming years by simplifying logistics of vaccine administration, improving supplies and reducing vaccination costs.^31^ Achieving the other two targets by 2030 will be a daunting task especially in the LMICs where millions of women have not received prophylactic vaccine, and the coverage of once in a lifetime screening is abysmally low.^32^

The landscape of cervical cancer screening is expected to change rapidly in the coming years with widespread introduction of HPV tests. Therapeutic vaccines may have a more meaningful role in the women detected to have high-risk HPV without any detectable lesions. Current standard-of-care for these women is frequent follow ups (in higher resourced settings) or ablative/excisional treatment (in resource-limited settings). The recent WHO recommendation to treat all high-risk HPV positive women with ablation or excision (after triaging if they are HIV positive) in the LMICs may not be feasible and sustainable, especially in countries with high HPV prevalence in the population.^8^ The observed high efficacy of therapeutic vaccines against high-risk HPV infection is encouraging, since the proportion of women with high-risk HPV clearing the infection following vaccination is double the proportion of women achieving spontaneous clearance. The excellent safety profile of the therapeutic vaccines reported in our systematic review gives more credence to this possibility.

A therapeutic vaccine with reasonably high efficacy to treat CIN 2/3 may have a high relevance in the LMICS where access to excision treatment facility is extremely limited and significant dropouts happen when women are referred to higher centers for treatment.^33^ Since excision treatment is significantly associated with preterm birth, low birthweight and premature rupture of membrane, a safer, simpler and less expensive alternative like therapeutic vaccination is likely to have a major public health impact.^34^ Unfortunately, except the MVA E2 vaccine none of the other vaccines available at present demonstrated sufficiently high efficacy to replace ablative or excisional treatment.

Many of the patients with CIN 2/3 have persistent high-risk HPV infection even after treatment with ablation or excision and they remain at a higher risk of recurrence.^35^ A systematic review has documented this proportion to be as high as 21% after 6 months.^36^ Concurrent administration of therapeutic vaccine during ablative or excisional treatment of high-grade lesions has been shown to induce higher viral clearance, which is likely to reduce the risk of recurrence.^25^ The vaccine induced increased CD8^+^ infiltrates are likely to contain HPV specific effector T cells establishing tissue residence in normal mucosa – leading to significantly less frequent recurrence and a long lasting protection.^16^ This is particularly important as women treated for high-grade cervical lesions remain at a high risk of developing cervical cancer or other anogenital cancers several years following treatment.^37,38^ Further studies are required to establish the role of therapeutic vaccines as an adjuvant to ablation or excision while treating CIN 2/3 lesions.

Our systematic review has certain limitations. We had to combine studies that were variable in case selection (e.g., some included CIN 2/3 lesions that were positive for HPV 16/18 only while others included lesions associated with any high-risk HPV types), ascertainment of endpoint (e.g., some studies included CIN 1 as evidence of regression while others didn’t), study designs (RCT, case-control studies or single-arm trials) and follow up protocols. Some of the studies did not have a control arm, which made it difficult to ascertain whether the responses observed were due to natural regression alone. Due to the limited number of phase II/III trials publishing their data till date we had no option but to combine their results.

The strength of our study is inclusion of only those studies that recruited histopathology proved untreated CIN 2/3 lesions and had rigorous criteria for ascertainment of disease at follow-up. This permitted us to provide a robust estimate of the efficacy of therapeutic vaccines against the most important endpoint of regression of high-grade lesions. A recently published systematic review of therapeutic vaccines showed that the overall incidence rate of CIN2/3 regression induced by the five types of vaccine was 62.48% [95% CI (42.80, 80.41)]. However, this estimate may be overestimated by including in their analysis both prophylactic and therapeutic vaccines, as the two vaccines have entirely different modes of action.^39^

To conclude, the modest efficacy of the therapeutic vaccines in the treatment of high-grade cervical cancer precursors may not justify replacing the highly effective ablative or excisional treatment with these new interventions. The existing vaccines may be made more effective through innovative approaches for treatment such as use of peptide-based vaccines to boost the effect of virus-based vaccines to overcome the anti-vector immunity.^9^ The possibilities of using the vaccines in HPV positive women to achieve a more rapid and durable clearance or as an adjunct to treating CIN 2/3 lesions with ablation or excision need to be explored further. The implementation issues like feasibility, acceptability, adoption, reach, cost and sustainability need to be studied in the contexts of both high and limited resourced settings.

## Data Availability

The related data for the current study are available from the corresponding author on reasonable request.

## Acknowledgments

We thank Teresa Lee and Latifa Bouanzi (IARC/WHO) for assistance in the design of search strategy terms and Krittika Guinot (IARC/WHO) for her help in the manuscript preparation and submission.

## Contributors

PB had the initial idea for the study. AIK, LZ developed the search strategy. AIK and LZ participated in the title and abstract screening, full-text screening and information extraction. PB consolidated the criteria based on the extracted information. RM assisted the data analysis. AIK, LZ, CS and PB drafted the manuscript and all authors provided critical comments on the manuscript. AIK and LZ have accessed and verified the data. All authors approved the final version of the manuscript. The corresponding author had final responsibility to submit for publication.

## Disclaimer

Where authors are identified as personnel of the International Agency for Research on Cancer / World Health Organization, the authors alone are responsible for the views expressed in this article and they do not necessarily represent the decisions, policy or views of the International Agency for Research on Cancer / World Health Organization.

## Funding

None.

## Completing interests

Authors declare no competing interests.

## Patient and public involvement

Patients and/or the public were not involved in the design, or conduct, or reporting, or dissemination plans of this research.

## Patient consent for publication

Not applicable.

## Provenance and peer review

Not commissioned.

## Supporting information

**Table S1.** Search strings for all four databases

| Pubmed database |  |
| --- | --- |
| # | Search |
| 1 | Papillomaviridae [MeSH Terms] OR papillomaviridae [Tiab] "human papilloma virus infection"[Tiab] OR "human papillomavirus infection"[Tiab] OR HPV[Tiab] OR "Human papillomaviruses"[Tiab] |
| 2 | "Cancer Vaccines"[MeSH] OR Therapeutic HPV vaccine* [Tiab] OR Immunomodulatory Therapy*[tiab] OR Injection Therapeutic Vaccine*[tiab] OR Therapeutic vaccine*[tiab] OR Vaccinotherapy*[tiab] OR therapeutic use[tiab] OR drug therapy[tiab] |
| 3 | "Randomized controlled trial"[pt] OR "Randomized controlled trials as topic"[mesh] OR "random allocation"[mesh] OR "double-blind method"[mesh] OR "single-blind method"[mesh] OR random*[tw] OR "Placebos"[Mesh] OR placebo[tiab] OR ((singl*[tw] OR doubl*[tw] OR trebl*[tw] OR tripl*[tw]) AND (mask*[tw] OR blind*[tw] OR dumm*[tw])) OR Clinical Trial, Phase II[pt] OR Clinical Trial, Phase III [pt] |
| 4 | #1 AND #2 AND #3 |

| Web of Sciences database |  |
| --- | --- |
| # | Search |
| 1 | Papillomaviridae OR papillomaviridae "human papilloma virus infection" OR "human papillomavirus infection" OR hpv OR "Human papillomaviruses" |
| 2 | "Papillomavirus Vaccines" OR "Cancer Vaccines" OR "Therapeutic HPV vaccines" OR Immunomodulatory Therapy* OR Therapies Immunomodulatory* OR Injection Therapeutic Vaccine* OR Vaccinotherapy* OR Therapeutic vaccine* |
| 3 | (Randomized Controlled Trial* near/1 (method* OR stud*)) OR (Controlled Clinical Trial near/1 stud*) |
| 4 | #1 AND #2 AND #3 |

| Embase database |  |
| --- | --- |
| # | Search |
| 1 | 'Papillomaviridae'/exp OR 'papillomaviridae':ti,ab,kw OR 'human papilloma virus infection':ti,ab,kw OR 'human papillomavirus infection':ti,ab,kw OR hpv:ti,ab,kw OR 'Human papillomaviruses':ti,ab,kw |
| 2 | 'Papillomavirus Vaccines' OR 'Cancer Vaccines' OR 'Therapeutic HPV vaccines' OR 'Injection Therapeutic Vaccine' OR Vaccinotherapy* |
| 3 | 'Randomized Controlled Trial' |
|  | #1 AND #2 AND #3 |
| 4 | #4 AND [embase]/lim NOT ('systematic review*' OR review*) NOT ('letter'/it OR 'editorial'/it OR 'note'/it) |

| Global Index Medicus (GIM) |  |
| --- | --- |
| # | Search |
| 1 | (Human papilloma virus infection) OR (human papillomavirus infection) OR HPV OR (Human papillomaviruses) |
| 2 | (Papillomavirus Vaccines) OR (Cancer Vaccines) OR (Therapeutic HPV vaccines) OR (Injection Therapeutic Vaccine) OR Vaccinotherapy* |
| 3 | (Randomized Controlled Trial) OR (Controlled clinical trial) |
|  | #1 AND #2 AND #3 |
| 4 | WPRIM (Western Pacific) (3) |

**Cochrane Central Register of Controlled Trials (Central)**
| # | Search |
| --- | --- |
| 1 | Papillomaviridae OR human papilloma virus infection OR HPV OR "human papillomavirus infection" OR "Human papillomaviruses" |
| 2 | Therapeutic HPV vaccines OR Vaccinotherapy OR Injection Therapeutic Vaccine |
| 3 | Randomized controlled trials as topic OR single-blind method OR Clinical Trial, Phase II OR Clinical Trial, Phase III |
|  | #1 AND #2 AND #3 |
| 4 | Cochrane Reviews (18)<br>Cochrane Protocols (1)<br>Trials (47) |

**Figure S1.**
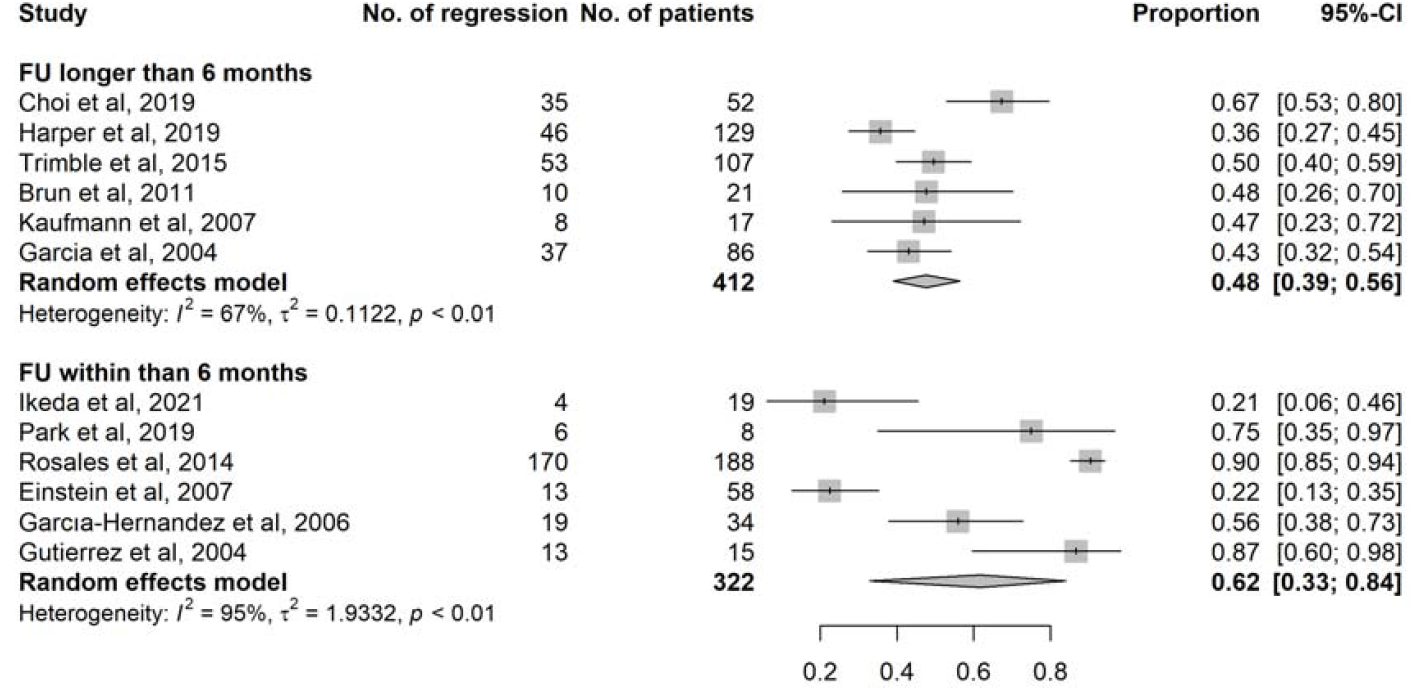
Proportion of CIN 2/3 lesions regressing to normal/CIN1 in vaccinated group by follow-up (FU) time (longer than 6 months vs. within 6 months)

**Figure S2.**
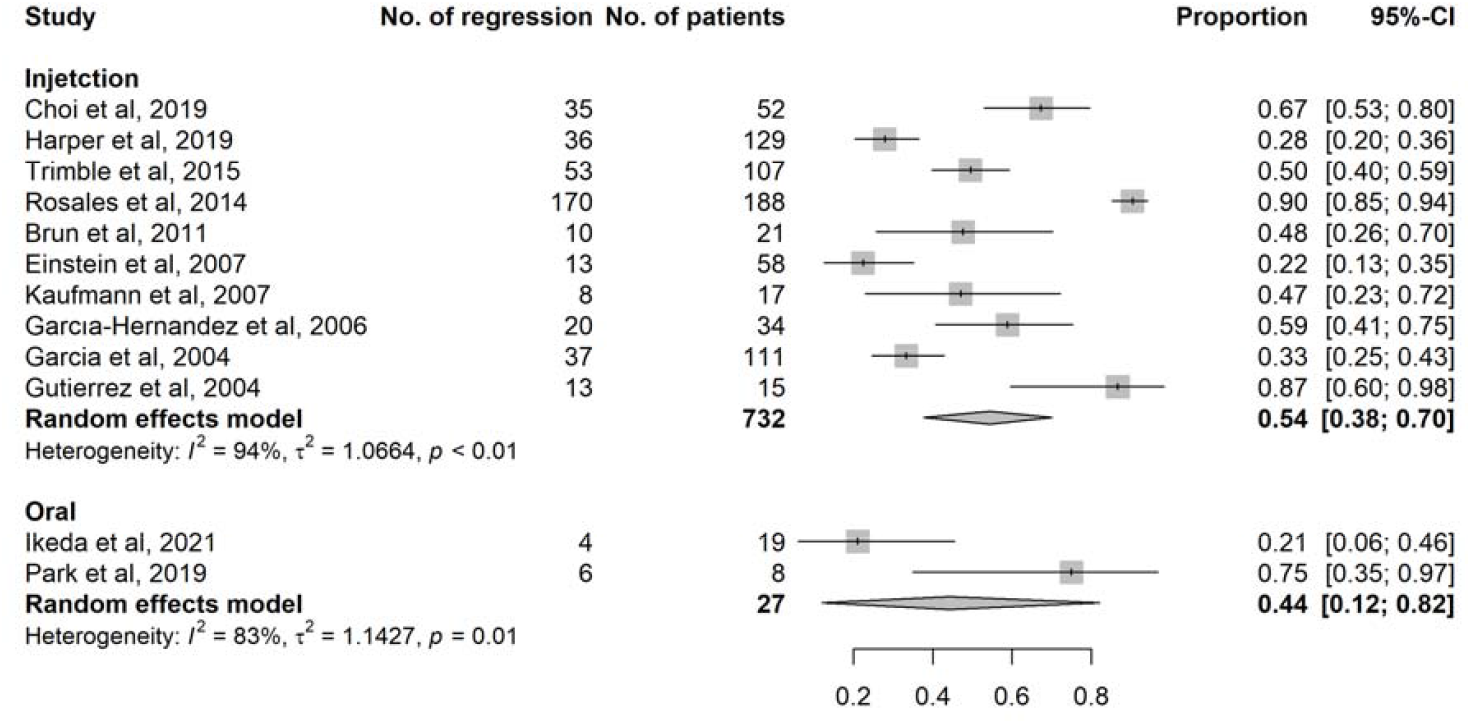
Proportion of CIN 2/3 lesions regressing to normal/CIN1 in vaccinated group by administration type (injectable vs. oral)

## Notes

### Competing Interest Statement

The authors have declared no competing interest.

### Funding Statement

This study did not receive any funding

## References

1. Loke AY, Kwan ML, Wong YT, Wong AKY. The Uptake of Human Papillomavirus Vaccination and Its Associated Factors Among Adolescents: A Systematic Review. J Prim Care Community Health. 2017; 8: 349–62.

2. Gravitt PE. The known unknowns of HPV natural history. J Clin Invest. 2011; 121: 4593–9.

3. Serrano B, Brotons M, Bosch FX, Bruni L. Epidemiology and burden of HPV-related disease. Best Pract Res Clin Obstet Gynaecol. 2018; 47: 14–26.

4. Guan P, Howell-Jones R, Li N, et al. Human papillomavirus types in 115,789 HPV-positive women: a meta-analysis from cervical infection to cancer. Int J Cancer. 2012; 131: 2349–59.

5. Petry KU. Management options for cervical intraepithelial neoplasia. Best Pract Res Clin Obstet Gynaecol. 2011; 25: 641–51.

6. Zhang L, Sauvaget C, Mosquera I, Basu P. Efficacy, acceptability and safety of ablative versus excisional procedure in the treatment of histologically confirmed CIN2/3: A systematic review BJOG. 2022; 10.1111/1471-0528.17251.

7. World Health Organization. WHO guidelines for the use of thermal ablation for cervical pre-cancer lesions. WHO: Geneva, 2019.

8. Bagarazzi ML, Yan J, Morrow MP, et al. Immunotherapy against HPV16/18 generates potent TH1 and cytotoxic cellular immune responses. Sci Transl Med. 2012; 4: 155ra38.

9. Smalley Rumfield C, Roller N, Pellom ST, Schlom J, Jochems C. Therapeutic Vaccines for HPV-Associated Malignancies. Immunotargets Ther. 2020; 9: 167–200.

10. Johnson LG, Armstrong A, Joyce CM, Teitelman AM, Buttenheim AM. Implementation strategies to improve cervical cancer prevention in sub-Saharan Africa: a systematic review. Implement Sci. 2018; 13: 28.

11. Ismaiel A, Ashfaq MZ, Leucuta DC, et al. Chemerin Levels in Acute Coronary Syndrome: Systematic Review and Meta-Analysis. Lab Med. 2022: lmac059.

12. Tufanaru C, Munn Z, Stephenson M, Aromataris E. Fixed or random effects meta-analysis? Common methodological issues in systematic reviews of effectiveness. Int J Evid Based Healthc. 2015; 13: 196–207.

13. Bell A, Fairbrother M, Jones K. Fixed and random effects models: making an informed choice. Quality & Quantity. 2019; 53: 1051–74.

14. Ikeda Y, Adachi K, Tomio K, et al. A Placebo-Controlled, Double-Blind Randomized (Phase IIB) Trial of Oral Administration with HPV16 E7-Expressing Lactobacillus, GLBL101c, for the Treatment of Cervical Intraepithelial Neoplasia Grade 2 (CIN2). Vaccines (Basel). 2021; 9: 329.

15. Harper DM, Nieminen P, Donders G, et al. The efficacy and safety of Tipapkinogen Sovacivec therapeutic HPV vaccine in cervical intraepithelial neoplasia grades 2 and 3: Randomized controlled phase II trial with 2.5□years of follow-up. Gynecol Oncol. 2019; 153: 521–9.

16. Trimble CL, Morrow MP, Kraynyak KA, et al. Safety, efficacy, and immunogenicity of VGX-3100, a therapeutic synthetic DNA vaccine targeting human papillomavirus 16 and 18 E6 and E7 proteins for cervical intraepithelial neoplasia 2/3: a randomised, double-blind, placebo-controlled phase 2b trial. Lancet. 2015; 386: 2078–88.

17. Garcia F, Petry KU, Muderspach L, et al. ZYC101a for treatment of high-grade cervical intraepithelial neoplasia: a randomized controlled trial. Obstet Gynecol. 2004; 103: 317–26.

18. Choi YJ, Hur SY, Kim TJ, et al. A Phase II, Prospective, Randomized, Multicenter, Open-Label Study of GX-188E, an HPV DNA Vaccine, in Patients with Cervical Intraepithelial Neoplasia 3. Clin Cancer Res. 2020; 26: 1616–23.

19. Kaufmann AM, Nieland JD, Jochmus I, et al. Vaccination trial with HPV16 L1E7 chimeric virus-like particles in women suffering from high grade cervical intraepithelial neoplasia (CIN 2/3). Int J Cancer. 2007; 121: 2794–800.

20. Rosales R, López-Contreras M, Rosales C, et al. Regression of human papillomavirus intraepithelial lesions is induced by MVA E2 therapeutic vaccine. Hum Gene Ther. 2014; 25: 1035–49.

21. García-Hernández E, González-Sánchez JL, Andrade-Manzano A, et al. Regression of papilloma high-grade lesions (CIN 2 and CIN 3) is stimulated by therapeutic vaccination with MVA E2 recombinant vaccine. Cancer Gene Ther. 2006; 13: 592–7.

22. Corona Gutierrez CM, Tinoco A, Navarro T, et al. Therapeutic vaccination with MVA E2 can eliminate precancerous lesions (CIN 1, CIN 2, and CIN 3) associated with infection by oncogenic human papillomavirus. Hum Gene Ther. 2004; 15: 421–31.

23. Park YC, Ouh YT, Sung MH, et al. A phase 1/2a, dose-escalation, safety and preliminary efficacy study of oral therapeutic vaccine in subjects with cervical intraepithelial neoplasia 3. J Gynecol Oncol. 2019; 30: e88.

24. Brun JL, Dalstein V, Leveque J, et al. Regression of high-grade cervical intraepithelial neoplasia with TG4001 targeted immunotherapy. Am J Obstet Gynecol. 2011; 204: 169.e1-8.

25. Einstein MH, Kadish AS, Burk RD, et al. Heat shock fusion protein-based immunotherapy for treatment of cervical intraepithelial neoplasia III. Gynecol Oncol. 2007; 106: 453–60.

26. Yeo-Teh NSL, Ito Y, Jha S. High-Risk Human Papillomaviral Oncogenes E6 and E7 Target Key Cellular Pathways to Achieve Oncogenesis. Int J Mol Sci. 2018; 19: 1706.

27. Arbyn M, Redman CWE, Verdoodt F, et al. Incomplete excision of cervical precancer as a predictor of treatment failure: a systematic review and meta-analysis. Lancet Oncol. 2017; 18: 1665–79.

28. Boilesen DR, Nielsen KN, Holst PJ. Novel Antigenic Targets of HPV Therapeutic Vaccines. Vaccines (Basel). 2021; 9: 1262.

29. WHO. Global strategy to accelerate the elimination of cervical cancer as a public health problem. Global strategy. World Health Organization: Geneva, 2020.

30. Bruni L, Saura-Lázaro A, Montoliu A, et al. HPV vaccination introduction worldwide and WHO and UNICEF estimates of national HPV immunization coverage 2010-2019. Prev Med. 2021; 144: 106399.

31. WHO. Highlights from the Meeting of the Strategic Advisory Group of Experts (SAGE) on Immunization 2022. Available from: https://cdn.who.int/media/docs/default-source/immunization/sage/sage-pages/sage_april2022meetinghighlights_11apr2022_final.pdf?sfvrsn=c2bd9f68_1

32. Lemp JM, De Neve JW, Bussmann H, et al. Lifetime Prevalence of Cervical Cancer Screening in 55 Low-and Middle-Income Countries. JAMA. 2020; 324: 1532–1542.

33. Selmouni F, Sauvaget C, Dangbemey DP, et al. Lessons learnt from pilot cervical cancer screening and treatment programmes integrated to routine primary health care services in Benin, Cote d’Ivoire and Senegal. JCO Global Oncology. 2022 (In press)

34. Kyrgiou M, Koliopoulos G, Martin-Hirsch P, Arbyn M, Prendiville W, Paraskevaidis E. Obstetric outcomes after conservative treatment for intraepithelial or early invasive cervical lesions: systematic review and meta-analysis. Lancet. 2006; 367: 489–98.

35. Castle PE, Schiffman M, Bratti MC, et al. A population-based study of vaginal human papillomavirus infection in hysterectomized women. J Infect Dis. 2004; 190: 458–67.

36. Hoffman SR, L. T, Lockhart A, et al. Patterns of persistent HPV infection after treatment for cervical intraepithelial neoplasia (CIN): A systematic review. Int J Cancer. 2017; 141: 8–23.

37. Ebisch RMF, Rutten DWE, IntHout J, et al. Long-Lasting Increased Risk of Human Papillomavirus-Related Carcinomas and Premalignancies After Cervical Intraepithelial Neoplasia Grade 3: A Population-Based Cohort Study. J Clin Oncol. 2017; 35: 2542–50.

38. Swedish KA, Factor SH, Goldstone SE. Prevention of recurrent high-grade anal neoplasia with quadrivalent human papillomavirus vaccination of men who have sex with men: a nonconcurrent cohort study. Clin Infect Dis. 2012; 54: 891–8.

39. Cai S, Tan X, Miao K, et al. Effectiveness and Safety of Therapeutic Vaccines for Precancerous Cervical Lesions: A Systematic Review and Meta-Analysis. Front Oncol. 2022; 12: 918331.

